# Detection of SARS-CoV-2 variant Mu, Beta, Gamma, Lambda, Delta, Alpha, and Omicron in wastewater settled solids using mutation-specific assays is associated with regional detection of variants in clinical samples

**DOI:** 10.1101/2022.01.17.22269439

**Authors:** Marlene Wolfe, Bridgette Hughes, Dorothea Duong, Vikram Chan-Herur, Krista R. Wigginton, Bradley J. White, Alexandria B. Boehm

## Abstract

Changes in the circulation of SARS-CoV-2 variants of concern (VOCs) may require changes in public health response to the COVID-19 pandemic, as they have the potential to evade vaccines and pharmaceutical interventions and may be more transmissive relative to other SARS-CoV-2 variants. As such, it is essential to track and prevent their spread in susceptible communities.We developed digital RT-PCR assays for mutations characteristic of VOCs and used them to quantify those mutations in wastewater settled solids samples collected from a publicly owned treatment works (POTW) during different phases of the COVID-19 pandemic. Wastewater concentrations of single mutations characteristic to each VOC, normalized by the concentration of a conserved SARS-CoV-2 N gene, correlate to regional estimates of the proportion of clinical infections caused by each VOC. These results suggest targeted RT-PCR assays can be used to detect variants circulating in communities and inform public health response to the pandemic.

**Importance:** Wastewater represents a pooled biological sample of the contributing community and thus a resource of assessing community health. Here we show that emergence, spread, and disappearance of SARS-CoV-2 infections caused by variants of concern are reflected in the presence of variant genomic RNA in wastewater settled solids. This work highlights an important public health use case for wastewater.

## Introduction

During an infectious disease outbreak it is critical to detect cases quickly and estimate the extent and timing of the outbreak to target interventions to mitigate spread. The detection of targets associated with infectious agents in wastewater can be used to infer information on the health of an entire population and provide critical outbreak monitoring services. This technique has been used widely during the COVID-19 pandemic, as SARS-CoV-2 RNA is readily detectable in wastewater and concentrations of RNA correlate to laboratory-confirmed COVID-19 infections in the contributing communities (1–4). Wastewater has previously been used to track gastrointestinal infections including poliovirus (5), and this work has extended to not only track COVID-19 (6) but also for other respiratory viruses such as respiratory syncytial virus (RSV) (7). Using wastewater to track community health has the advantage of providing information on an entire community without relying on individual clinical testing, which may be expensive or unavailable and requires individuals to alter their behavior to seek testing. Wastewater may be a leading indicator of community health when shedding by infectious individuals precedes symptom onset.

The COVID-19 pandemic has seen SARS-CoV-2 acquire mutations that have given rise to variants with distinguishing characteristics. Variants of concern (VOCs) or interest (VOIs) are variants that may evade vaccines or other pharmaceutical interventions, be more transmissible, or cause more severe illness. Variant classifications by the World Health Organization (WHO) and the US Centers for Disease Control and Prevention (CDC) have changed over the course of the pandemic, but VOCs are named according to the Greek alphabet and have included Alpha, Beta, Gamma, Delta, Lambda, Mu, and Omicron (8, 9). The emergence of variants is primarily identified by sequencing of clinical specimens; this same approach is then typically used to track the spread of VOCs into and throughout communities. A health department or clinical laboratory will choose a subset of all specimens to sequence, and results are usually available within two weeks. This data could lack community representation if samples from some clinics are more likely to be sequenced than others, or may be biased when specific samples are chosen for sequencing because of patient characteristics. A two week processing time may prevent a fast public health response to a spreading variant of concern.

Monitoring variants in wastewater may overcome some of the problems with relying on sequencing clinical specimens to track variant emergence and spread. A wastewater sample is representative of the entire contributing community and therefore lacks bias that is common for sequencing of clinical specimens. However, a wastewater sample is more complex than a clinical specimen: it contains many different types of viruses (10) that have undergone different degrees of degradation (11). Sequencing SARS-CoV-2 RNA from wastewater likely requires enrichment or amplification of the SARS-CoV-2 genome (12). An alternative approach for variant tracking in wastewater is application of targeted RT-PCR assays that amplify and allow detection of short genomic sequences characteristic to the variant.

Several publications to date have explored the use of targeted assays to detect SARS-CoV-2 variants in wastewater. Heijan et al. (13) applied a commercial digital RT-PCR assay to wastewater influent samples to detect a single nucleotide polymorphism (SNP) (mutation N501Y) present Beta and Alpha. Lee et al. (14) and Graber et al. (15) applied RT-QPCR assays that detect mutations present in Alpha to wastewater samples. Yaniv et al. developed RT-QPCR assays for Gamma and Delta (16), and Alpha and Beta (17) and applied them to four or 10 wastewater samples, respectively, as proof of concept. To date, there is limited research (18, 19) to apply targeted assays for characteristic mutations of diverse variants to wastewater samples across different phases of the pandemic to identify emergence patterns, and compare those to data from variants in clinical specimens.

The present study develops novel targeted digital droplet (dd-)RT-PCR assays for the detection of six characteristic mutations from distinct variants in wastewater. In particular, we develop and utilize assays for mutations characteristic of Alpha, Beta & Gamma, Delta, Mu, Lambda, and Omicron and then measure these in wastewater solids from a publicly owned treatment work (POTW) located in the Bay Area of California, USA. We measure concentrations in wastewater settled solids as concentrations of SARS-CoV-2 RNA are enriched several orders of magnitude in solids relative to liquid wastewater (20, 21). We subsequently compare the measurements to data on occurrence of those variants in clinical specimens, aggregated at the state-level.

## Materials and methods

### Assay Development

Assays were designed to target mutations characteristic of the following variants: Alpha (HV69-70), Delta (del156-157/R158G), Beta & Gamma (together) (E484K/N501Y), Mu (del256-257), Lambda (del247-253), and Omicron (del143-145) (Table 1). These characteristic mutations were chosen because they are present in high percentages of the associated variant sequences in GISAID (Table 1, information accessed through outbreak.info), and they represent deletions or multiple single nucleotide polymorphisms (SNPs) in close proximity and thus are likely to be more specific than assays targeting a SNP. Assays were developed in silico using Primer3Plus (https://primer3plus.com/). Mutation and adjacent sequences were obtained from genomes downloaded from NCBI. The parameters used in assay development (that controlled sequence length, GC content, and melt temperatures) are provided in Table S1. Primers and probe sequences are provided in Table 2. The development and testing of the HV69-70 and del156-157/R158G are reported elsewhere (19), so additional details are not provided on these assays herein.

**Table 1.** Variants included in this study (column 1), the characteristic mutations that ddRT-PCR assays were developed for (column 2), the percent of variant genomes with the mutation(s) in column 2 (column 3), the positive control used in the sensitivity testing experiments (column 4), and the SARS-CoV-2 genomes that were used, along with the respiratory panel, in the specificity testing conducted in vitro (column 4).

| Variant Name(s) | Mutation(s) (gene location) | Percentage of variant genomes, observed globally, with mutation(s) in GISAID as of 10 Jan 2022 (total number of variant genomes with mutations / total number of variant genomes) | Positive control used in sensitivity testing | SARS-CoV-2 genomes tested against *in vitro* for specificity |
| --- | --- | --- | --- | --- |
| Alpha | HV69-70 (del69-70) (S gene) | 97% (1,106,137 /1,143,476) | Reported in Yu et al. (19) | Reported in Yu et al. (19) |
| Delta | del156-157/R158G (S gene) | 92% (3,644,016 /3,953,372) | Reported in Yu et al. (19) | Reported in Yu et al. (19) |
| Beta & Gamma | E484K/N501Y (S gene) | Gamma: 94% (112925/119761)<br>Beta: 85% (34576/40553) | Positive clinical swab sequenced as P.1 (Gamma) | WT gRNA and Alpha |
| Mu | del256-257 (ORF3a) | 95% (13,978 /14,712) | Positive clinical swab from Stanford sequenced as B.1.621 | WT gRNA, Alpha, Beta, Delta and Gamma |
| Lambda | del247-253 (S gene) | 84% (8029/9577) | gRNA from cultivated Lambda variant from Pinsky Lab C.37 | WT gRNA, Alpha, Beta, Gamma, Delta, Mu |
| Omicron | del143-145 (S gene) | 95% (212,997/224,673) | Synthetic Omicron gRNA from Twist control 48 | WT gRNA, Alpha, Beta, Gamma, Delta, Mu, Lambda |

**Table 2.**
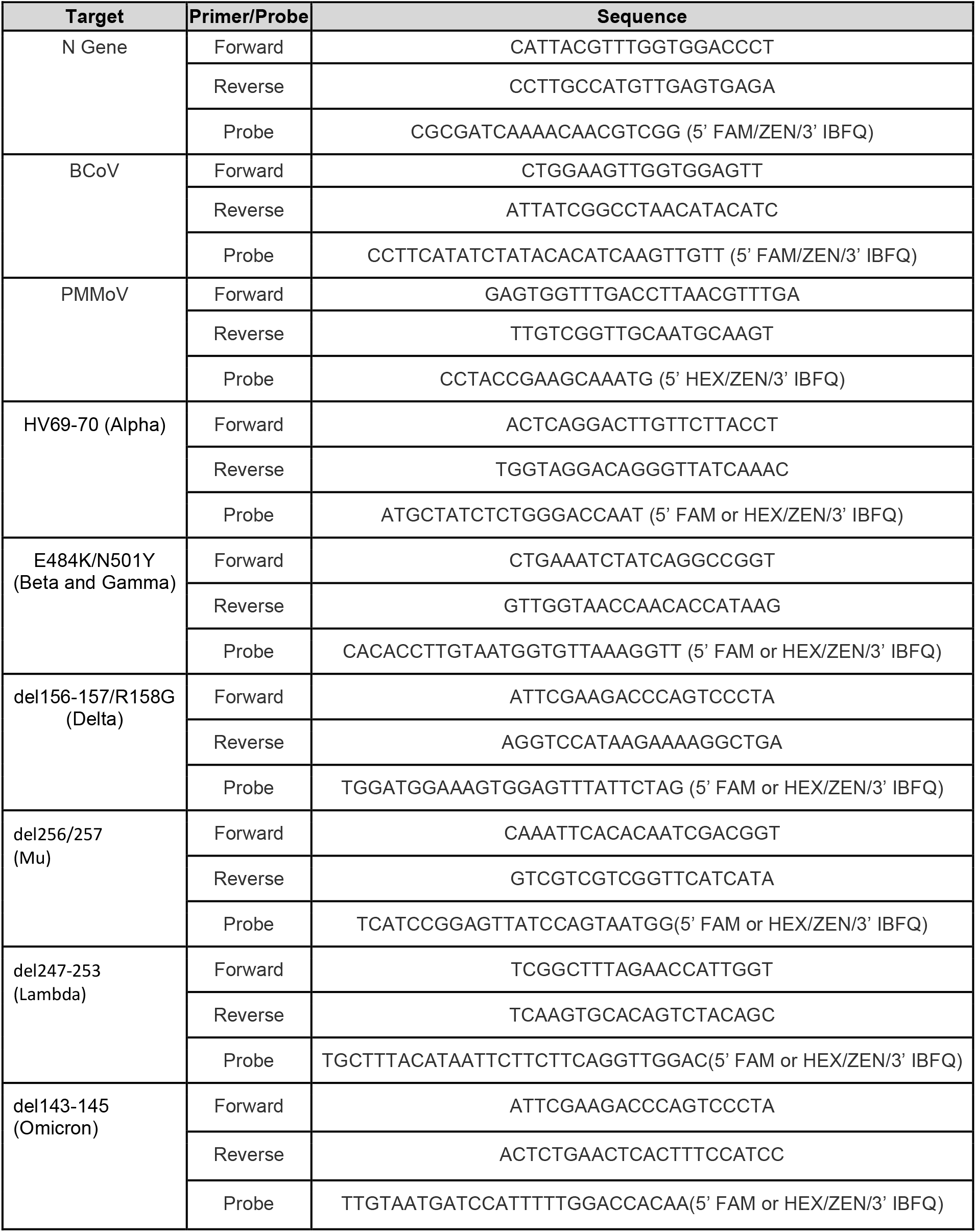
Primer and probe sequences used in this study to target characteristic mutations in variants. The variant containing the characteristic mutation is shown below the name of the targeted mutation.

### Specificity Screening Against Other Targets

Primers and probe sequences were screened for specificity in silico using NCBI Blast, and then tested in vitro against a virus panel (NATtrol™ Respiratory Verification Panel, NATRVP2-BIO, Zeptomatrix) that includes several influenza and coronavirus viruses, “wild-type” gRNA from SARS-CoV-2 strain 2019-nCoV/USA-WA1/2020 (ATCC® VR-1986D™) which does not contain the mutations (hereafter referred to as WT-gRNA) as well as a combination of heat inactivated SARS-CoV-2 stain B.1.1.7 (SARS-CoV-2 variant B.1.1.7, ATCC® VR-3326HK™), a positive clinical sample confirmed as Mu provided by Dr. Ben Pinsky at Stanford Virology Laboratory, and synthetic gRNA from Twist Biosciences (South San Francisco, California, USA) for Beta (Twist control 16), Gamma (Twist control 17), Delta (Twist control 23), and Omicron (Twist control 48) (Table 1). RNA was extracted from the virus panel and whole viruses using the Perkin Elmer Chemagic Viral RNA extraction kit (Chemagic Kit CMG-1033-S designed for SARS-CoV-2). RNA was used undiluted as template in digital droplet PCR with mutation primer and probes (see further details on digital PCR below). The concentration of targets used in the in vitro specificity testing was approximately 275 copies per well. The mutation assays were challenged against the respiratory panel gRNA in single wells, and non-target variant gRNA in 8 replicate wells. Positive PCR controls (Table 1) were included on each plate.

The sensitivity and specificity of the mutation assays were further tested by diluting target variant gRNA (Table 1) for the mutations in no (0 copies), low (100 copies), and high (10,000 copies) background of WT-gRNA. Each dilution was run in three replicate wells. The number of copies of variant mutation sequences input to each well was estimated using a dilution series of variant gRNA in no background; the vendor specified concentration of the variant gRNA was scaled by the slope of the curve relating the measured ddRT-PCR concentration and the calculated input concentration based on the vendor estimates. Our experience suggests vendor estimates can be imprecise. PCR negative controls were run in 4 wells per plate.

### Wastewater samples

A publicly owned treatment work (POTW) that serves populations in Santa Clara County, California, USA (San José-Santa Clara Regional Wastewater Facility) was included in the study. It serves approximately 1,500,000 people; further description of the POTW can be found in Wolfe et al. (1).

Samples of approximately 50 mL of settled solids were collected by POTW staff using sterile technique in clean, labeled bottles. POTW staff manually collected a 24 h composite sample (21). Samples were immediately stored at 4°C, transported to the lab, and processed within 6 hours of collection.

Samples were collected daily for a larger COVID-19 wastewater surveillance effort starting in November 2020 (1), and a subset of these samples are used in the present study and were chosen to span the period prior to and including presumed emergence of different variants. Generally, sampling was about once per week or month prior to presumed emergence and then 3-7 times per week during and after the period of emergence. Details on sampling frequency are provided in Table 3. A previous study (19) reported Alpha mutation data for the POTW and those data are included in our analysis for completeness. That same study reported some Delta mutation data (N = 48, data until 1 Aug 2021) for the POTW and that data are included here. The methods below describe those used for the new measurements including those for Mu, Lambda, Beta/Gamma, Delta (measured daily between 1 Aug 2021 and 2 Jan 2022), and Omicron mutations.

**Table 3.**
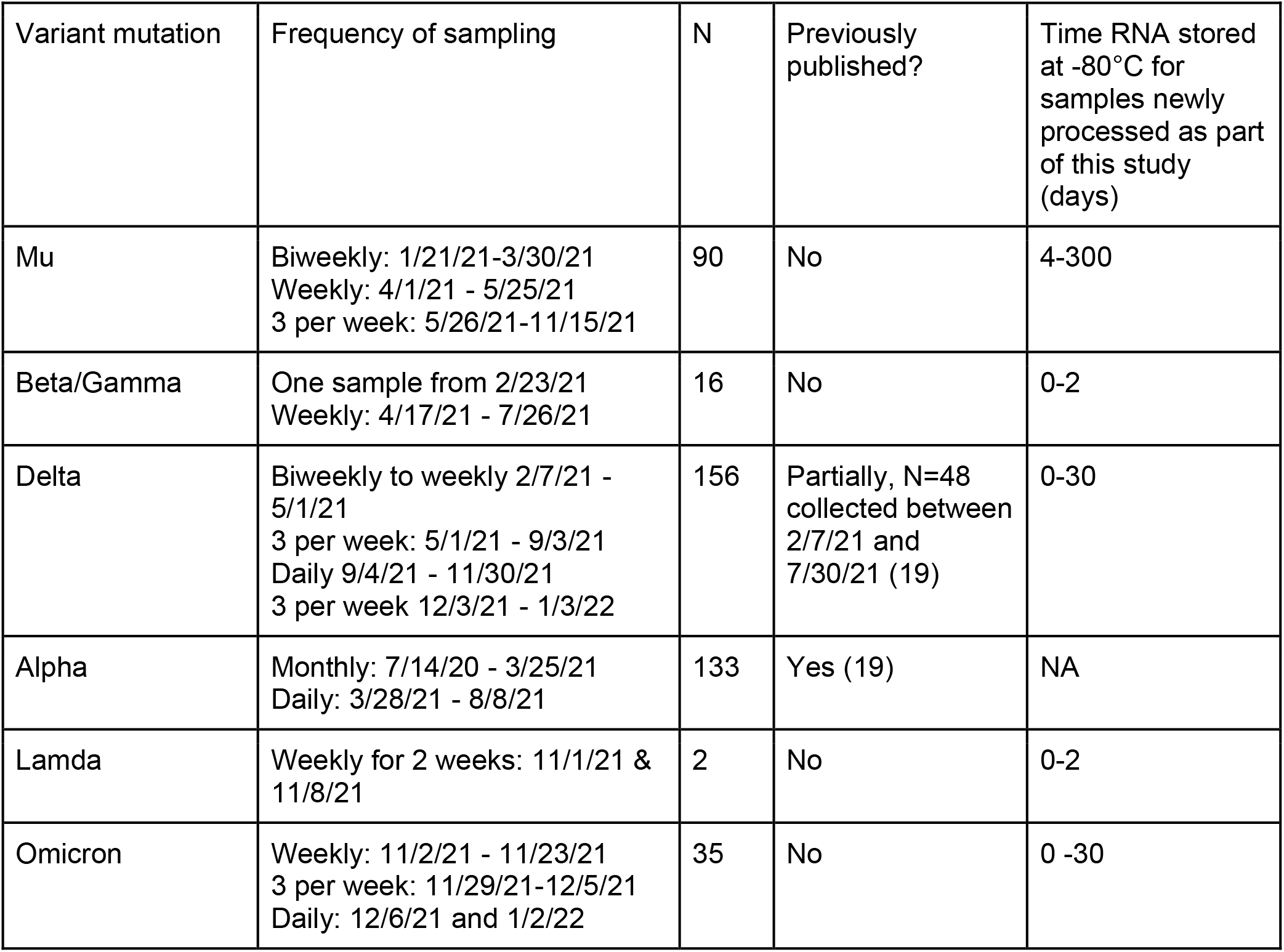
Frequency of sample collection for different assay applications, number of samples included in this study, whether any of the data have been published, and the time range that RNA samples were stored between extraction of RNA and running the PCR assays. RNA extraction occurred on the day of sample collection, as explained in the methods.

RNA was extracted from the 10 replicate aliquots of dewatered settled solids as described elsewhere (1, 22, 23). This process includes dilution of the solids in DNA/RNA Shield (Zymo, Irvine, CA) as a means to alleviate inhibition (24). RNA was subsequently processed immediately (within 24 h of sample collection) to measure concentrations of the N gene of SARS-CoV-2, pepper mild mottle virus (PMMoV), and bovine coronavirus (BCoV) recovery using digital droplet RT-PCR methods described in detail elsewhere (1, 25). The N gene assay targets a region of the N gene that is conserved across these variants. PMMoV is highly abundant in human stool and wastewater globally (26, 27) and is used as an internal recovery and fecal strength control for the wastewater samples (28). BCoV was spiked into the samples and used as an additional recovery control; all samples were required to have greater than 10% BCoV recovery. RNA extraction and PCR negative and positive controls were included to ensure no contamination as described in Wolfe et al. (1) The N gene measurement was multiplexed with the Delta mutation assay in samples processed after Aug 1, 2021, and the Omicron mutation assay in samples processed after 6 Dec 2021. For the other mutation assays, the extracted RNA was stored at -80°C for a period of time (Table 3) before it was analyzed for the N gene and the Mu, Beta/Gamma, or Lambda mutation assay in a multiplex digital RT-PCR assay. The SARS-CoV-2 N gene was run a second time for assays run on stored RNA to test for RNA degradation during storage at -80°C (no to minimal degradation was observed, see supporting material, SM). Each of the 10 replicate RNA extracted were run in its own well, and the 10 wells were merged for analysis. Wastewater data are available publicly at the Stanford Digital Repository (https://purl.stanford.edu/hs561fr5902); results below are reported as suggested in the EMMI guidelines for reporting ddRT-PCR measurements in environmental samples (29).

### ddRT-PCR

Digital RT-PCR was performed on 20 μl samples from a 22 μl reaction volume, prepared using 5.5 μl template, mixed with 5.5 μl of One-Step RT-ddPCR Advanced Kit for Probes (Bio-Rad 1863021), 2.2 μl Reverse Transcriptase, 1.1 μl DTT and primers and probes at a final concentration of 900 nM and 250 nM respectively. Template was diluted 1:100 for measuring PMMoV and BCoV. Primer and probes were purchased from IDT (sequences in Table 3). Droplets were generated using the AutoDG Automated Droplet Generator (Bio-Rad). PCR was performed using Mastercycler Pro with the following cycling conditions: reverse transcription at 50°C for 60 minutes, enzyme activation at 95°C for 5 minutes, 40 cycles of denaturation at 95°C for 30 seconds and annealing and extension at 61°C (for SARS-CoV-2 targets) or 56°C (for PMMoV/BCoV targets) for 30 seconds, enzyme deactivation at 98°C for 10 minutes then an indefinite hold at 4°C. The ramp rate for temperature changes were set to 2°C/second and the final hold at 4°C was performed for a minimum of 30 minutes to allow the droplets to stabilize. Droplets were analyzed using the QX200 Droplet Reader (Bio-Rad). All liquid transfers were performed using the Agilent Bravo (Agilent Technologies).

Thresholding was carried out using QuantaSoft™ Analysis Pro Software (Bio-Rad, version 1.0.596). In order for a sample to be recorded as positive, it had to have at least 3 positive droplets.

For the wastewater samples, concentrations of RNA targets were converted to concentrations per dry weight of solids in units of copies/g dry weight using dimensional analysis. The dry weight of the dewatered solids was determined by drying (23). Using this approach, three positive droplets corresponds to a concentration between ∼500-1000 cp/g; the range in values is a result of the range in the equivalent mass of dry solids added to the wells. The total error is reported as standard deviations and includes the errors associated with the Poisson distribution and the variability among the 10 replicate wells.

### Variants present in regional clinical specimens

The 7-d, centered, rolling average fraction of clinical specimens sequenced from the State of California classified as Alpha, Beta, Gamma, Mu, Lambda, Delta, and Omicron as a function of specimen collection data were acquired through outbreak.info which collates data from GISAID. Data were downloaded from outbreak.info on January 5, 2022 for all variants, except for Omicron for which data were downloaded on January 10, 2022. Data were acquired in the form of time series plots, and data were extracted using PlotDigitizer (https://plotdigitizer.com/).

### Statistics

We hypothesize that wastewater concentrations of characteristic variant mutations are associated positively with the proportion of infections caused by the variant in the contributing population. Because data on incidence rates of COVID-19 caused by specific variants at the sewershed level are not readily available, we used state-level data on the fraction of sequenced clinical specimens identified as specific variants to represent this variable. We normalized the wastewater concentration of the variant mutation by the concentration of the N gene to represent the fraction of total SARS-CoV-2 RNA (represented by the N gene assay target which is conserved across variants) that comes from the variant; hereafter this concentration is referred to as the relative concentration of the mutation. We applied a five adjacent sample box-average smoothing algorithm to the relative concentrations to aid in visualization, but used raw data in statistical analyses. We used Kendall’s tau (hereafter tau) to test for associations between the relative concentration and the fraction of clinical specimens assigned to the corresponding variant as the two variables were not normally distributed (Shaprio Wilk test, p<0.05 for all). The measured relative concentration was matched to the 7-d, centered, rolling average fraction of clinical specimens classified as the associated variant obtained from outbreak.info.

## Results

### Lambda, Mu and Beta/Gamma Mutation Assay specificity

In silico analysis of the Lambda, Mu Beta/Gamma, and Omicron mutation assays indicated no cross reactivity between the variant mutation assays and deposited sequences in NCBI. When challenged against the respiratory virus panel and gRNA from WT-SARS-CoV-2 and other variants (Table 1) no cross reactivity was observed. When mutation assays were tested on their target variant gRNA diluted in a background of high and low WT-SARS-CoV-2 RNA, there was no evidence of cross reactivity (Figure 1). Positive and negative controls run on all the ddPCR plates were positive and negative. These results suggest that the variant mutation ddRT-PCR assays are specific. Yu et al. (19) provide details on the specificity and sensitivity of the Alpha and Delta mutation assays, which are also specific and sensitive.

**Figure 1.**
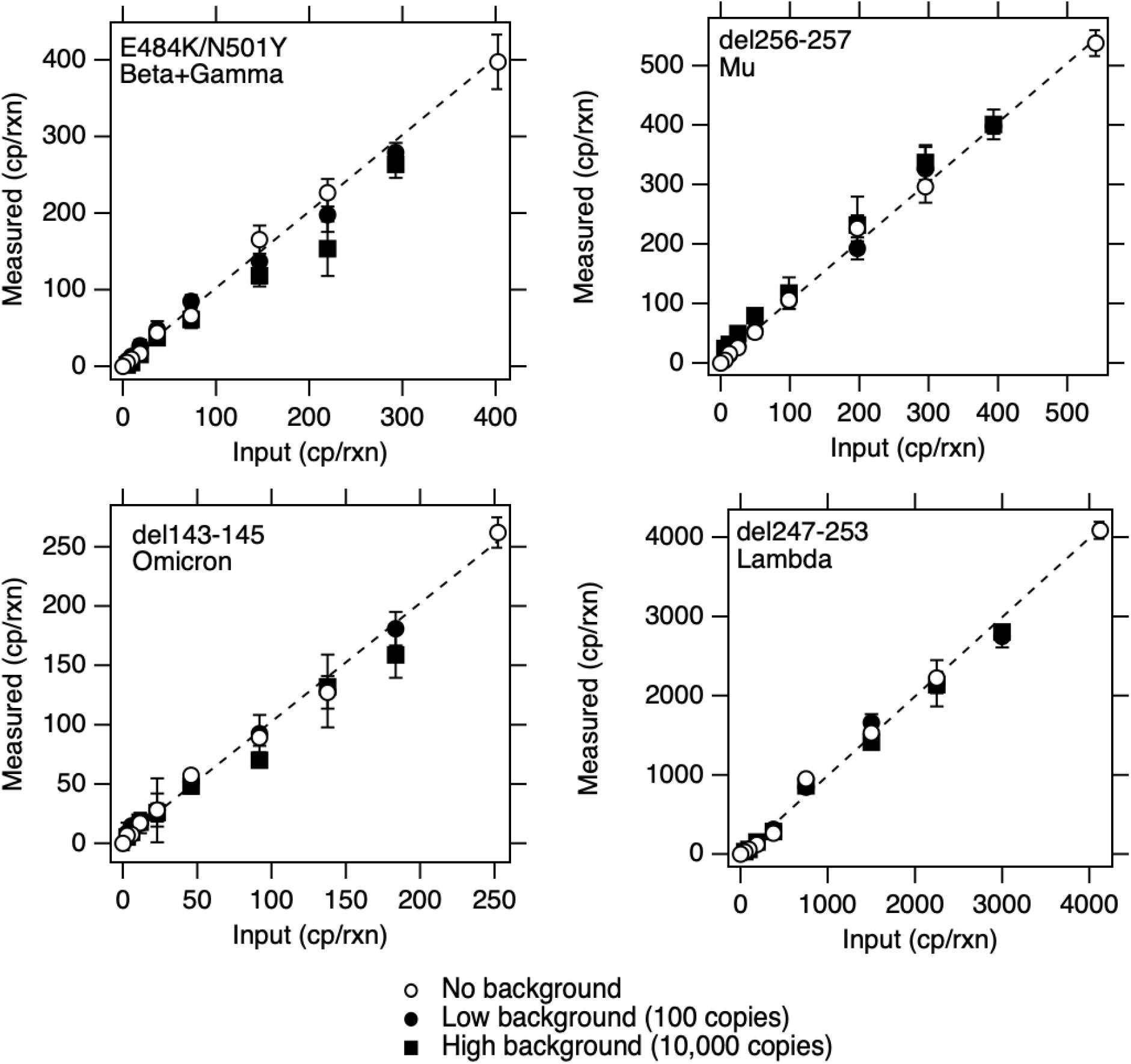
Copies (cp) of mutations measured when RNA containing the mutation was diluted into no, low, and high background of WT-gRNA. Low background is 100 copies/well and high background is 10,000 copies/well where “copies” refers to copies of genomes of WT-gRNA. Markers show average cross three replicate wells and error bars represent standard deviations. In some cases, the error bar is not visible because it is smaller than the marker.

### Variant mutation RNA in wastewater

All positive and negative controls were positive and negative respectively, indicating assays performed well and without contamination. BcoV recoveries were higher than 10% and PMMoV concentrations were within the expected range for the POTW suggesting an efficient and acceptable recovery of RNA during RNA extraction (Figure S1).

As described previously by Yu et al. (19), the Alpha mutation was not detected in wastewater solids prior to January 2021. After that time, it was detected in low relative concentrations until late March 2021, when its relative concentration started to increase until early June 2021 when its relative concentration peaked. The concentration began to decrease until the mutation became undetectable in late June 2021 (Figure 2).

**Figure 2.**
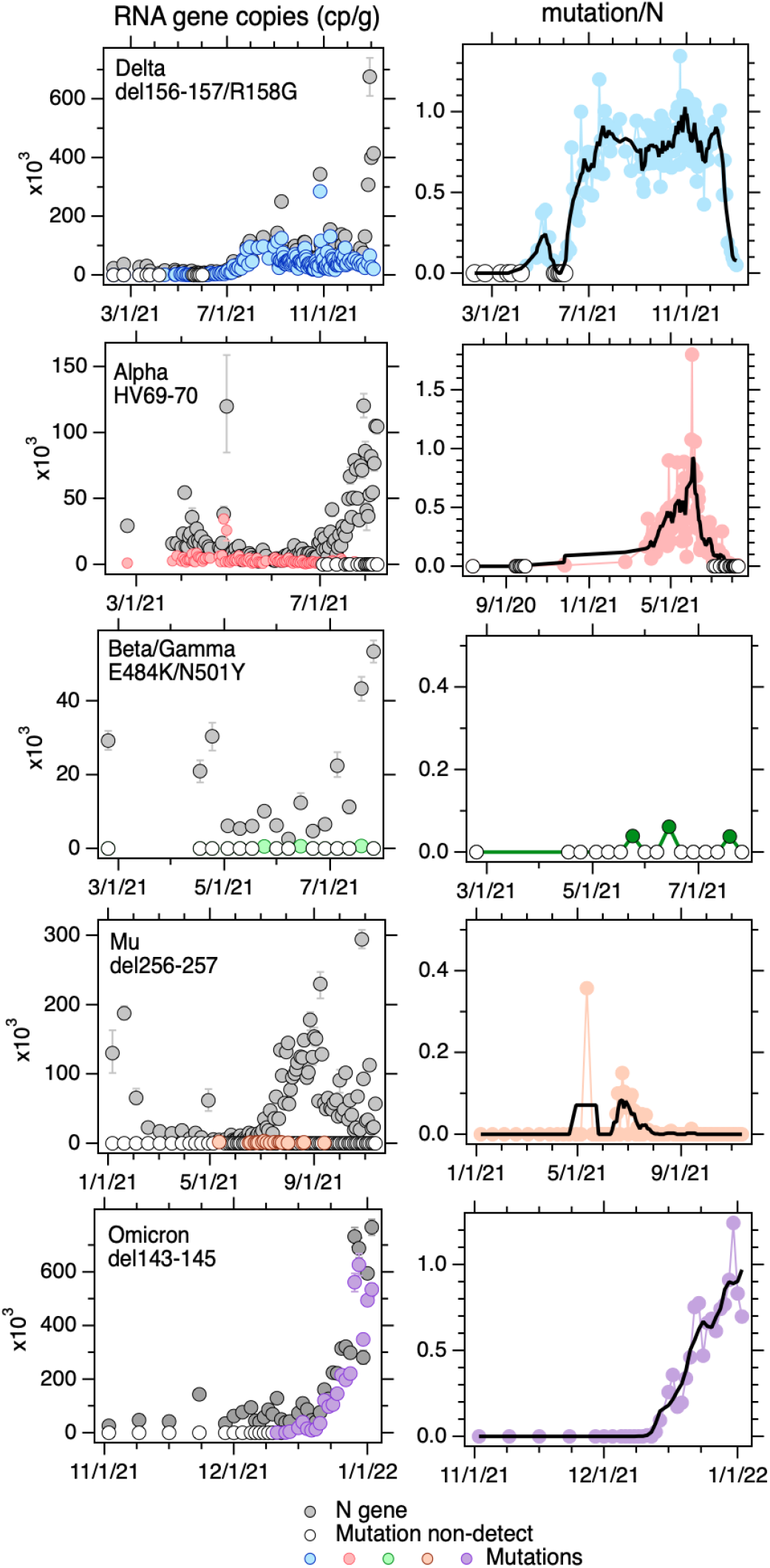
Left column. Concentrations (copies (cp) per gram dry weight) of the N gene and the indicated mutation in wastewater solids as a function of time. Open circles indicate non-detects for the mutation gene. Error bars represent standard deviations and include poisson error and replicate well error and was output from the ddPCR machine software as “total error”. Right column. The concentration of the mutation normalized by the concentration of the N gene as a function of time (“relative mutation concentration”, unitless). The black line represents the 5-point smoothed value for the dates. Open circles are non-detects. Non-detects are shown as 0 on the plots. The Alpha mutation data are from Yu et al. (19); Delta mutation data through 7/31/21 are from Yu et al. (19).

The delta mutation was not detected in wastewater solids until early April 2021 at which time it increased and was detectable for about a month before it fell to non-detectable levels again for two weeks. Thereafter, the concentration of the delta mutation rose over the month of June until it was present at about the same concentration as the N gene, thereafter the concentration stayed approximately equivalent to the N gene until the beginning of December 2021 (Figure 2). The relative concentration subsequently decreased until the end of December 2021.

The omicron mutation was absent in the samples tested prior to 11 December 2021. After first detected on 11 December, the concentrations rose steadily until the relative concentration was close to 1 at the end of December (Figure 2).

The mutation present in Beta and Gamma was rarely detected in wastewater solids (Figure 2). It was not detected in wastewater until late May 2021 when it was detected at a very low relative concentration. It was detected a total of 3 times between late May 2021 and the end of July 2021, all at low concentrations relative to the N gene.

The mutation present in Mu was not detected until May 2021 when it was detected at a fairly high concentration relative to the N gene in a single sample. Thereafter, it was not detected again until mid June after which its relative concentration increased for 1 month until the beginning of July and then decreased over the following month until the beginning of August after which the mutation was no longer detected (Figure 2). The lambda mutation assay was applied to 2 samples in November 2021 and was not detected.

The 5-sample smoothed relative concentrations of the Alpha, Delta, Mu, and Omicron mutations, and the raw relative concentrations of the Beta/Gamma mutations are shown in Figure 3 along with the 7-d rolling average fraction of clinical specimens from the state assigned as each variant. The temporal trends in the relative wastewater concentrations and clinical specimen data are qualitatively similar. The wastewater variant mutation data (raw data, Figure 2) are positively, significantly associated with the clinical variant data (tau = 0.75, p<10^−15^ for Alpha, tau = 0.42, p<10^−13^ for Delta, tau= 0.91, p<10^−13^ for Omicron, tau = 0.36, p<10^−4^ for Mu) with the exception of data for Beta/Gamma. The relative concentration of the Beta/Gamma mutation was positively associated with the fraction of clinical specimens assigned as Beta and Gamma (tau = 0.14, p = 0.5), but the association was not statistically significant. This may be due to the relatively low cadence of measurements as we only measured the mutation once per week; this is low compared to frequency of variability typically observed in wastewater measurements (1). There was no reported case of Lambda in the state from November 2021, and our lack of detection of the Lambda mutation in that month is consistent with this. The positive associations between relative variant mutation concentrations and the fraction of clinical specimens assigned to Alpha and Delta is consistent with findings described by Yu et al. (19) using sewershed-aggregated clinical data over a different time period.

**Figure 3.**
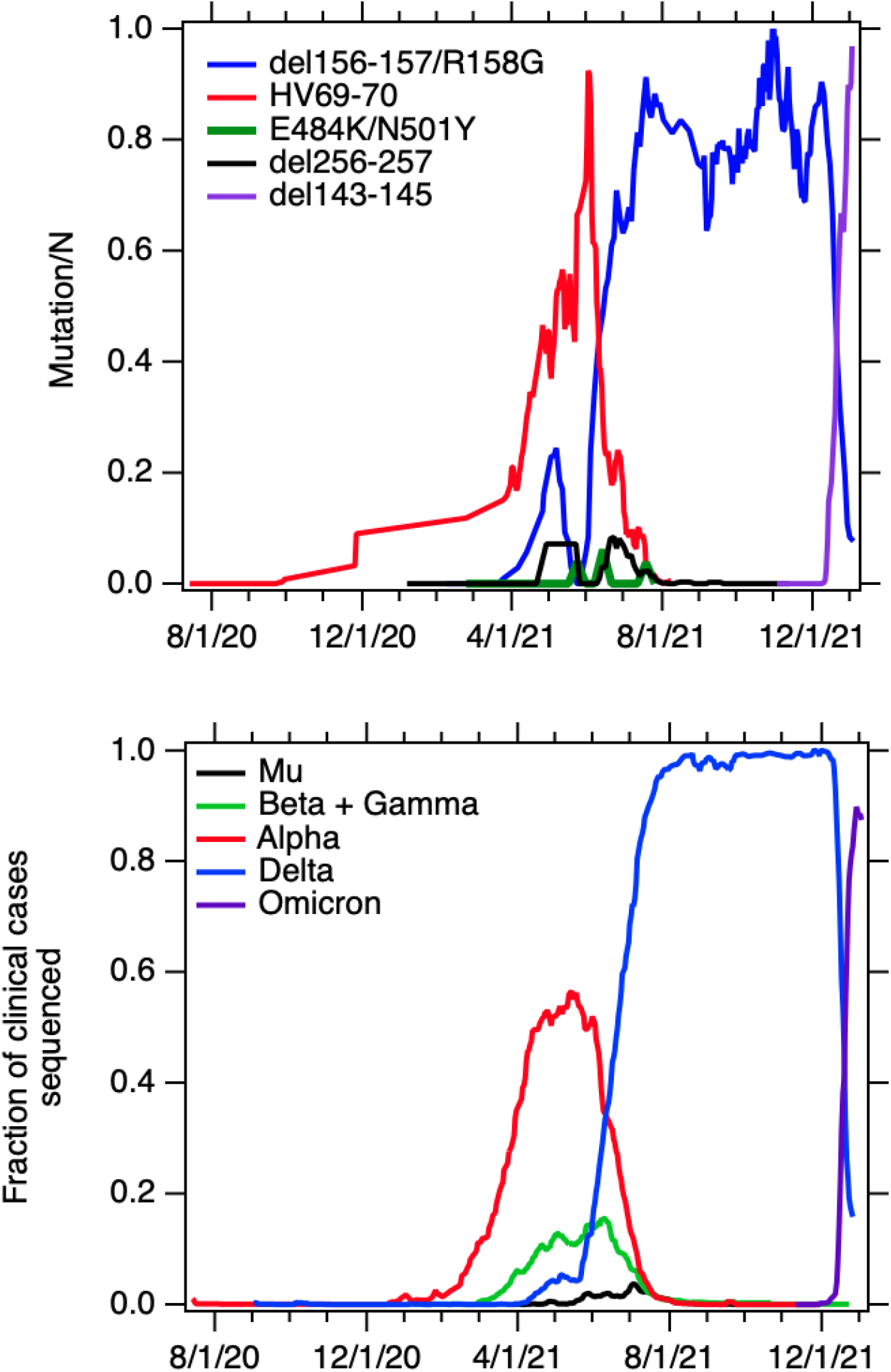
Top graph. Five-point smoothed relative concentrations of mutations in wastewater solids (unitless) with the exception of E484K/N501Y which is the raw data, non-detects were taken as 0. Bottom plot. The fraction of all sequenced clinical specimens in California that were classified as the indicated variant (7-d rolling average from outbreak.info).

## Discussion

Wastewater results are indicative of the replacements of consecutive variants in circulation over time. The decline in relative wastewater concentrations of the Alpha mutation is coincident with the rise in relative wastewater concentrations of the Delta mutation suggestive of Delta outcompeting Alpha in causing infections in susceptible populations. Beta and Gamma mutations began to appear in wastewater along with Mu mutations as the relative concentration of the Delta mutation was rising. It appears these variants were also present but not able to compete with Delta as their relative concentrations decreased to non-detect shortly after their appearance in wastewater. The increase in relative wastewater concentrations of the Omicron mutation is coincident with the decline in the relative concentrations of the Delta mutation suggesting Omicron potentially outcompeting Delta, or a large increase in Omicron incident cases atop of a stable background of Delta incident cases.

Several other studies have reported agreement between detection of characteristic variant mutations in wastewater and the occurrence of variants in clinical specimens. Lee et al. (14) report a three fold higher increase in a characteristic mutation in Alpha in wastewater from January to March 2021, comparable to an increased fraction in Alpha sequences from clinical samples deposited in GISAID during the same time period. Graber et al. (15) report agreement in wastewater trends of a characteristic mutation from Alpha and data aggregated at the city level on Alpha circulation based on clinical specimens. Yaniv et al. (18) report lack of detection of a characteristic mutation in Alpha in wastewater when clinical data suggests it was not circulating.

The clinical data used in this study are imperfect. The fraction of sequenced clinical specimens assigned to each variant may be biased by the selection of specimens to sequence, and the number of specimens sent for sequencing. The data displayed at Figure 3 are aggregated across the state, and may not reflect the occurrence of infections caused by different variants in the population contributing to the sewershed and represented in the wastewater data, particularly for variants with low occurrence rates. Despite these limitations, the wastewater variant mutation measurements correlate well with the variant clinical data.

SARS-CoV-2 RNA in wastewater is a complex mixture of gRNA of all circulating variants in a given community. SARS-CoV-2 gRNA present in wastewater may be present in an intact or damaged viral capsid with or without an envelope (30), and may have undergone damage or fragmentation (11). In contrast, a clinical specimen contains numerous copies of one SARS-CoV-2 variant, with the gRNA likely intact. Given the complexity of wastewater SARS-CoV-2 gRNA, the presence of a single characteristic mutation in wastewater cannot definitively indicate that a variant is present because a variant is defined by the presence of multiple mutations on a single genome. A single characteristic mutation detected in a wastewater sample could theoretically be from a different variant, known or unknown, containing the same mutation. Even the detection of two mutations characteristic of a specific variant in wastewater does not prove the variant is present, because those two mutations could have originated from different genomes. Moreover, the characteristic mutations used in this study are not present in 100% of the associated variant genomes. Despite these limitations, our results suggest that the concentration of a single mutation characteristic of a variant of concern over the concentration of a conserved SARS-CoV-2 target (the N gene) is associated with the proportion of regional infections caused by the variant.

These findings suggest that for variants of concern, valuable insights are available on the circulation of the variants through the use of wastewater, and these insights are attainable using assays that target a single characteristic variant mutation. Development of assays for SARS-CoV-2 variants requires in silico assay design, procurement of primers, probes and positive control material, and specificity and sensitivity testing. The rate limiting step in this process, we have found, is the procurement process. Targeted ddRT-PCR assays can be applied to samples with a turnaround time for results of less than 24 hours, and new targeted assays can be quickly developed and applied to wastewater when new variants are identified and expected to spread into communities to gain insight into their local emergence. We were able to implement this process in real-time for development and implementation of the Omicron mutation assay, which we were able to apply to daily samples at this POTW starting 6 Dec 2021 to capture the emergence of the variant at high resolution.

## Data Availability

All data produced in the present work are contained in the manuscript

## Acknowledgments

This work was funded by a gift from the CDC Foundation. The funders had no role in study design, data collection and interpretation, or the decision to submit the work for publication. We acknowledge the following individuals for assistance with wastewater solids collection: Payal Sarkar, Noel Enoki, and Amy Wong.

